# Shared decision-making about uveal melanoma treatment in the Netherlands: non-neutral framing of medical information

**DOI:** 10.64898/2026.06.25.26356547

**Authors:** Aryana Shirzada, Lisa Vlug, Marina Marinkovic, Gré P.M. Luyten, Jaco C. Bleeker, T.H. Khanh Vu, Coen R.N. Rasch, Nanda Horeweg, Arwen H. Pieterse

**Author notes:** Corresponding author: Aryana Shirzada, Department of Radiation Oncology, Leiden University Medical Center (LUMC) Albinusdreef 2, 2333 ZA, Leiden, The Netherlands. This study was funded by The Dutch Cancer Society. The funding organization had no role in the design or conduct of this research. Grant number: 2021-13400.

## Abstract

**Background:** A subset of uveal melanomas can be treated using either enucleation or proton beam therapy (PBT), which offer similar oncological outcomes. The most appropriate treatment depends on a patient’s preference. To allow patients to genuinely determine their preference, it is recommended to describe options as neutrally as possible. This study assesses to what extent ocular oncologists use and perceive non-neutral framing behaviour, and if it is related to patient satisfaction with decision-making.

**Methods:** Consultations of ocular oncologists with patients newly-diagnosed with uveal melanoma were audio recorded, transcribed verbatim, and coded for ocular oncologists’ explicit and implicit non-neutral framing behaviours. Explicit non-neutral framing was defined as: explicitly mentioning a preferred option at least once, without relating it to the patient. Implicit non-neutral framing was defined as: describing an option (un)favourably, without providing a medically substantive clarification alongside.

**Results:** 110 patients provided consent for the audio recordings. Non-neutral framing was found in 84% (n=92/110) of consultations. We found explicit behaviour in 38% (42/110) and implicit behaviour in 76% (84/110, median=1, range, 0-4) of consultations. The most frequent implicit framing was presenting options by positively or negatively emphasizing one option. Non-neutral framing behaviours were not significantly related to patient satisfaction with decision-making.

**Conclusion:** This study shows that in most consultations some non-neutral framing was present, which did not impact patients’ satisfaction with decision-making. Nonetheless, ocular oncologists should be aware that how they describe options may influence preferences in ways that do not align with the patient’s values.

## 1. Introduction

A subset of choroidal melanomas can be treated by enucleation or using proton beam therapy (PBT) (1-3). The treatment options are associated with similar risks of metastases and mortality (4), but differ regarding other characteristics, including procedures, side effects (2, 5, 6), and impact on quality of life (7, 8). Given the comparable oncological benefits and differences on other aspects, the optimal treatment for a clinically eligible patient depends on their individual preferences. This makes shared decision-making (SDM) the most appropriate clinical decision-making model (9). As part of the SDM process, ocular oncologists will have to learn what the patient considers most important in relation to the available treatment options (10). Because the decision is novel, most patients lack immediate preferences and instead construct them as they learn about the options. Consequently, patients require relevant and sufficient information regarding the nature, procedures, and possible outcomes of the treatment options, in light of the natural course of the disease. To make a choice about treatment, the characteristics that distinguish the two treatments are most relevant. Our previous research has clarified which distinguishing characteristics are most important for newly-diagnosed patients who face the decision between enucleation and PBT, to be informed about (11). We also assessed to what extent ocular oncologists and radiation oncologists mention these characteristics routinely to patients facing the treatment choice (11). Our previous study did not assess how ocular oncologists communicate medical information to patients. The information ocular oncologists provide should support preference formation and therefore should ideally be conveyed as neutrally as possible, in accordance with current SDM models (10). Formulating information neutrally is a practice to strive for, and is expected to help patients in forming preferences that genuinely reflect their goals and priorities (12). In this study, we consider information provision not to be neutral, i.e., non-neutral, when two conditions are met: (i) the ocular oncologist presents medical information about a treatment option in ways that are favourable or unfavourable to that option, and (ii) this framing is unrelated to what a patient has made clear to consider important or to prefer, up to that point. Non-neutral framing can be explicit or implicit, both of which can imply a “better” choice. Research has shown that in real-life consultations, physicians often present information about treatment options in ways that go beyond factual descriptions and patient values are not considered (13-16). In simulated scenarios, patients tend to choose treatments contradicting their personal preferences when physicians frame treatment information non-neutrally (17). In addition, when patients are asked about SDM, they report that the physician’s opinion plays the most important role in decision-making (18), and therefore might influence the patient’s preferences. Importantly, in other simulated decision situations a treatment poorly aligned with patient preferences may result in less decision satisfaction, more decision regret, or suffering more from side-effects (17, 19).

This study aimed to assess a) to what extent and how often ocular oncologists frame treatment options non-neutrally, b) to what extent ocular oncologists and patients perceive that the oncologist has framed options non-neutrally, and c) whether non-neutral framing of treatment options is related to patient satisfaction about decision-making, among patients currently faced with the decision between enucleation and PBT.

## 2. Methods

### 2.1 Study design

This is a cross-sectional study using audio recordings and questionnaires to evaluate information provision during consultations in which ocular oncologists discuss treatment options with patients newly-diagnosed with uveal melanoma, and/or with ciliary body involvement. From June 2019 to April 2024, patients were included at the Leiden University Medical Center (LUMC), The Netherlands, which serves as the Dutch national referral centre for uveal melanoma. Following the initial intake and diagnostic testing, the ocular oncologist discloses the final diagnosis and informs the patient the treatment options. This consultation about final diagnosis and treatment was audio recorded for the study. Finally, patients meet with an oncology nurse for practical information, are referred to a radiation oncologist if PBT is considered, or are offered a follow-up telephone consultation if they wish more decision-making support.

At the start of the consultation, the ocular oncologist asked for verbal consent from patients to audio record the consultation. All participants provided written consent to participate in this study. This research was approved by the Medical Ethical Committee of the LUMC (P17.195) and was registered at www.clinicaltrials.gov (identifier: NTC05377957).

### 2.2 Participants

Adults above 18 years were eligible for inclusion if diagnosed with uveal melanoma that could be treated with either enucleation or PBT. Ruthenium brachytherapy could also be an option depending on tumour dimension or localisation. Patients with distant metastases at time of diagnosis, a local recurrence, or those eligible only for Ruthenium brachytherapy were excluded. All four practicing ocular oncologists (GL, JB, TV, MM) audio recorded consultations, with a small subset recorded by residents or ocular oncologist in training.

### 2.3 Coding of the consultations

All audio recordings were transcribed verbatim. We identified and coded any instance of non-neutral framing behaviour, and patient treatment-related preferences. *Explicit non-neutral framing* (present/absent) was defined as the ocular oncologist explicitly mentioning at least once his or her preferred option without explicitly relating the preference to the patient’s individual characteristics. For example, this could entail that the ocular oncologist did not explain that because a patient’s vision in the contralateral eye is reduced, PBT is preferred over enucleation, as PBT offers some probability of keeping vision in the affected eye. *Implicit non-neutral framing* (frequency) was defined as the ocular oncologist describing an option in a way that includes an evaluation and thereby puts the option in a favourable or unfavourable light, without providing a medically substantive clarification alongside. Implicit non-neutral framing can occur in various forms. We used an existing coding scheme (14) which covers four categories of non-neutral framing: (1) Persuading patients by using (*clinical*) expertise; (2) Creating the *illusion of decisional control*, by giving the patient a sense of involvement while the physician in fact makes the treatment decision; (3) Presenting treatment recommendations as *authorized decisions*, by presenting the treatment option as guideline-based; and (4) Presenting benefits and side-effects of the treatment options in *unbalanced ways*. From the original scheme we selected nine that could apply to our clinical context, across all four categories (Table 2; items described in Supplementary Table A). We added travel distance as an implicit non-neutral framing behaviour which is relevant since enucleation requires fewer follow-up consultations than PBT. Additionally, we coded the treatment favoured by each framing behaviour, if the patient expressed a treatment-related preference, and the timing of it, and whether a (preliminary) treatment decision was made. Treatment-related preference can encompass either statements from patients about how they evaluate aspects of the treatment options, or a treatment preference. Notably, final consultation preferences or decisions did not always correspond to the actual received treatment. The overall framing direction was determined by the explicit preference, or, if absent, the majority of implicit behaviour. Two assessors (AS and LV) coded all transcripts independently and resolved disagreements by consensus.

### 2.4 Patient and oncologist questionnaires

The patients and ocular oncologists, respectively, completed the iSHAREpatient or iSHAREphysician questionnaire (Supplementary Tables B and C) post-consultation (both) and pre-treatment (patients) (20). The iSHARE questionnaires include the same 15 items, but the items are mirrored. Patients completed the questionnaire once; the ocular oncologists completed it for each of their participating patients. The questionnaires cover six dimensions, of which the following three are most relevant to the present study: Choice awareness (2 items), Medical information (7 items), and Decision (1 item). These dimensions were selected to assess perspectives on information provision, the consideration of patient preferences, and whether the patient’s preferences had been incorporated in treatment decision-making.

The iSHARE questionnaires use 6-point scales ranging from 0 (not at all) to 5 (completely). Internal consistency of the Choice awareness and Medical information dimensions was assessed using Cronbach’s α, which showed good to excellent internal consistency among patients (α=0.84-0.90), and moderate to excellent internal consistency among ocular oncologists (α = 0.63-0.93). Dimension scores are calculated by averaging the scores on the relevant items. Higher dimension scores indicate more explicit clarification by the ocular oncologists that there is choice, and that patient preferences are relevant in making the decision (Choice awareness), more comprehensive and balanced information provision (Medical information), and decision-making that reflects the integration of patients’ perspective more (Decision).

In addition, patient satisfaction with decision-making was assessed using the 6-item Satisfaction With Decision scale (SWD) (21). Each item is scored on a 5-point scale from 1 (strongly disagree) to 5 (strongly agree), and total scores are computed by adding item scores (range total score, 6 – 30). The SWD scale showed excellent internal consistency (Cronbach’s α, 0.98). Higher scores indicate greater patient satisfaction with decision-making. Patient age and final treatment decision were collected from the electronic health record.

### 2.5 Statistical methods

Descriptive statistics were used to report participant characteristics. Median and mean scores are reported for the questionnaires. Spearman’s rank correlation coefficients (ρ) were computed to assess the associations between, respectively, the iSHAREpatient and iSHAREphysician dimensions and the presence or absence of explicit non-neutral framing behaviour, and number of implicit non-neutral framing behaviours. In addition, a linear mixed-effects model (LMM) was used to assess potential significant differences in correlations between non-neutral framing behaviour and scores on the three iSHAREphysician and iSHAREpatient dimensions across the four ocular oncologists. Spearman’s rho correlation analysis was further used to assess the association between SWD scores and the occurrence of explicit non-neutral framing behaviour, and the number of implicit non-neutral framing behaviours. Corresponding 95% confidence intervals (CI) were determined. P-values below 0.05 using a two-sided test were considered statistically significant. Correlations were interpreted according to the following cut-off values: 0.00-0.59, unacceptable; 0.50-0.59, poor; 0.60-0.69, questionable; 0.70-0.79, acceptable; 0.80-0.89, good; 0.90-1.00, excellent (22, 23). Statistical analysis was performed using SPSS Statistics v. 29.0 (IBM, Armonk, NY, USA).

## 3. Results

### 3.1 Participants

The four ocular oncologists included a total of N=110 patients (range, 18-43). Sixty-one (56%) patients were female, and their mean age was 64 years (SD = 11.8). The average duration of consultations was 13 minutes (SD = 5.1).

### 3.2 Observed non-neutral framing

We observed non-neutral framing in 92/110 (84%) consultations, and it was absent in 18/110 (16%) (Table 1). Overall, non-neutral framing was observed in favour of PBT in 54/92 (59%) consultations and in favour of enucleation in 24/92 (26%) consultations. In 15/92 (16%) consultations, both enucleation and PBT were supported by a variety of implicit non-neutral framing behaviours within the same consultation. The ocular oncologists framed treatment options in favour of PBT in 30-82% of consultations, and in favour of enucleation in 18–70%.

**Table 1.** Frequency and direction of non-neutral framing behaviours in 110 consultations.

|  |  | Total<br>N<sup>b</sup> (%) | In favour of which treatment: |  |  |
| --- | --- | --- | --- | --- | --- |
|  |  |  | Enucleation<br>N (%) | PBT<br>N (%) | Both<br>N (%) |
|  | *Total number of consultations in which explicit non-neutral framing behaviour occurred* | 42 (38)<sup>c</sup> | 6 (14)<sup>d</sup> | 36 (86)<sup>d</sup> | 0 (0) |
|  | *Total number of implicit non-neutral framing behaviours* | 84 (75)<sup>c</sup> | 47 (56)<sup>e</sup> | 22 (26)<sup>e</sup> | 15 (18)<sup>e</sup> |
| **Creating the illusion of decisional control** | 1. From mild to more serious treatment – a gradual decision | 44 (52) | 2 (5) | 42 (95) | 0 (0) |
| **Persuading patients using (clinical) expertise** | 2. Deterring vs encouraging: *using others as an example* | 32 (38) | 13 (40) | 3 (9) | 16 (50) |
|  | 3. Selectively presenting side-effects and/or benefits | 2 (2) | 1 (50) | 1 (50) | 0 (0) |
| **Unbalanced presentation of benefits and side-effects** | 4. Emphasizing benefits or side-effects of treatment | 25 (30) | 17 (68) | 6 (24) | 2 (8) |
|  | 5. Presenting side-effects after final treatment decision had been made | 10 (11) | 7 (75) | 3 (25) | 0 (0) |
|  | 6. Minimising the treatment's impact | 10 (11) | 5 (50) | 5 (50) | 0 (0) |
|  | **7.** Making assertions about the patient's personality or health status | 7 (8) | 2 (29) | 5 (71) | 0 (0) |
| **Presenting treatment recommendations as authorized decisions** | **8.** Presenting treatment as an authorized 'we' decision | 3 (3) | 2 (75) | 1 (25) | 0 (0) |
|  | **9.** Presenting treatment as an authorized decision based on 'the guideline' | 2 (2) | 1 (50) | 1 (50) | 0 (0) |
| **Logistics** | **10.** Travel distance | 5 (5) | 1 (20) | 4 (4) | 0 (0) |

Percentages in the "Total" column may exceed 100% as multiple forms of implicit non-neutral framing behaviour could occur per consultation.
<sup>a</sup> Six behaviours from the original coding scheme were excluded, because they focused specifically on adjuvant treatment in the context of primary breast cancer, and were therefore deemed not relevant (12).
<sup>b</sup> Percentages are calculated with respect to the total number of consultations in which non-neutral framing (also) occurred unless otherwise stated.
<sup>c</sup> Percentages are calculated with respect to the entire group
<sup>d</sup> Percentages are calculated with respect to the total number of consultations in which *explicit* non-neutral framing occurred (n = 42)
<sup>e</sup> Percentages are calculated with respect to the total number of consultations in which *only implicit* non-neutral framing occurred (n = 51)

Explicit non-neutral framing occurred in 42/110 (38%) consultations, and in 9/42 (21%) of these this was the only form of non-neutral framing that was observed. When explicit framing was used, this was usually in favour of PBT (n=36/42, 86%). For example: *“Both are possible. I think, I would, I do tend to go a bit towards the protons then, rather than directly removing the eye, but that’s really very personal*.*”*

Implicit non-neutral framing occurred in 84/110 (76%) consultations, and in 51/84 (60%) this was the only form of non-neutral framing. Overall, implicit non-neutral framing behaviours were observed with a median of 1.0 (range, 0–4) per consultation. Four instances were documented in three consultations. Phrases that were used to describe PBT in an implicit non-neutral manner were, e.g., *“It is a treatment with quite some hassle. It is called proton beam therapy”*. Presenting treatments in order of mild to more serious (n=43/84, 51%) occurred most often, such as by stating *“We have listed [the treatments] in order of attractiveness”* (Table 2). PBT was then most often mentioned first (n=41/43). Another common type of implicit non-neutral framing was using other people’s experiences regarding a specific treatment (31/84, 37%). Ocular oncologists would present PBT more favourably than enucleation by saying: *“Most people are more nuanced, and they say: if I lose all of my vision, I still retain my own eye, and that is already a lot*.*”* Similarly, ocular oncologists would present enucleation more favourably by saying for example: *“Someone else would say: all of those procedures [PBT], and follow-up consultations, and whatever, that is never going to have a good ending*.*”* One of the more strongly worded statements was: *“And other people say: well, if I lose my eye, life will be over for me. Then I’ll really… That’s not going to happen*.*”* In half of the consultations in which other people’s experiences were used to describe a treatment option, implicit non-neutral framing behaviour occurred towards both treatment options (n=16/31, 52%).

**Table 2.**
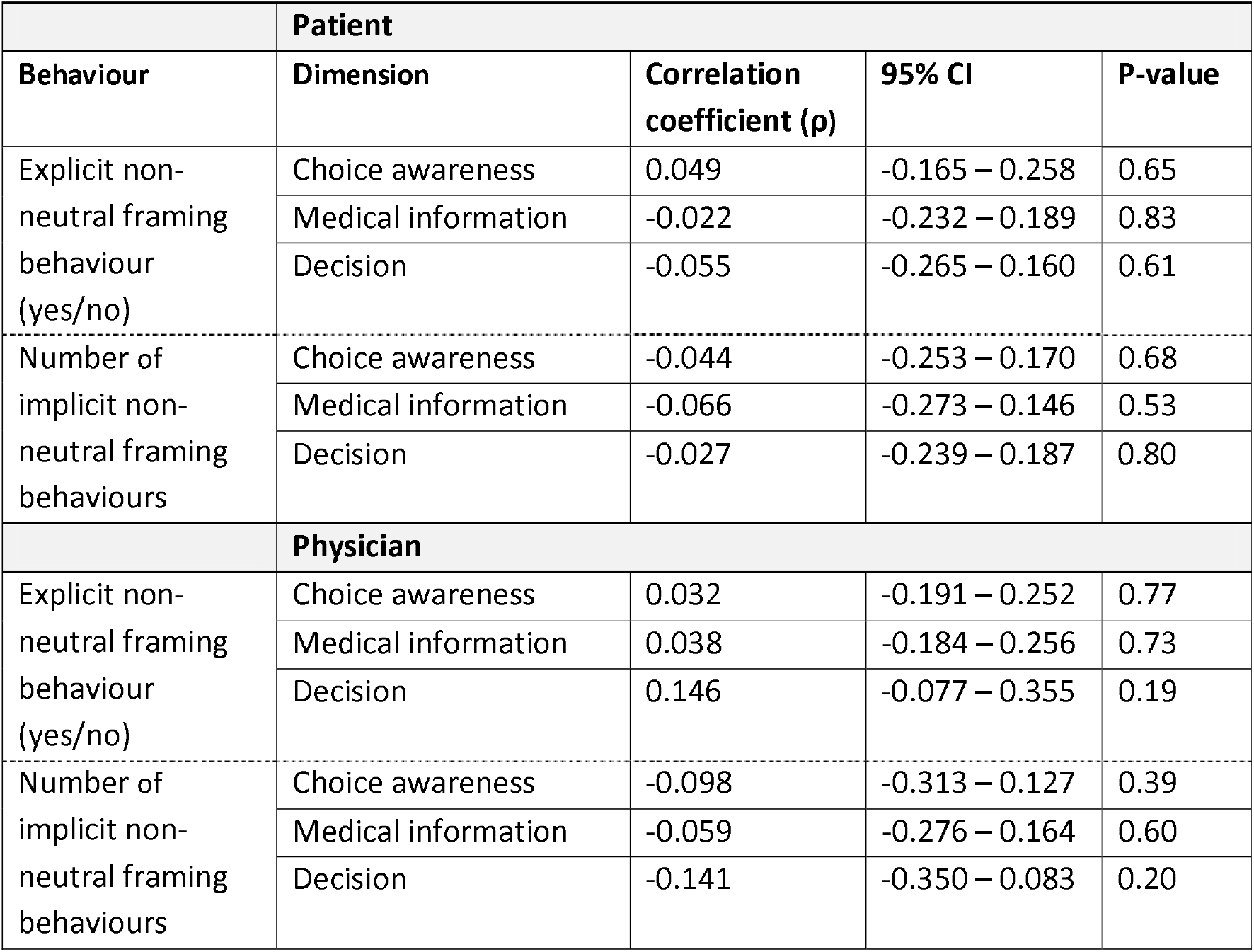
Correlations between non-neutral framing behaviours and the iSHARE dimensions (Choice awareness, Medical information, and Decision) in patients (N = 92) and ocular oncologists (N = 4)

In 33/84 (39%) consultations, either the possible benefits or side effects of the two treatment options were overly emphasized. For example, focusing on the possible burden of PBT by repeatedly describing the trajectory of PBT as lengthy and challenging. An example is: *“It is quite an intensive process, which means you will need to visit this hospital and the clinic in Delft several times, [*..*] I want to stress that proton therapy is a very demanding treatment*.[..*] Again, I must emphasize that this specific trajectory is particularly intensive*.*”*

In 5/84 (6%) consultations, the ocular oncologist mentioned travel distance in favour of a treatment option by saying, e.g. *“Proton therapy is a bit more complex, because we have to do an MRI before the surgery, and after the surgery, and you will have regular check-ups for which you need to come here. Well, fortunately, you don’t live so far from here*.*”* A list with examples of each implicit non-neutral framing behaviour can be found in Supplemental Table D.

### 3.3 Patient treatment preference

In total, 54/110 (49%) patients explicitly communicated a treatment preference during the consultation, and in 3/54 (6%) consultations, patients did so before the oncologist framed information in a non-neutral manner. Overall, patients stated a preference for PBT in 34/54 (63%), and for enucleation in 13/54 (24%) consultations. One patient said to prefer Ruthenium brachytherapy, which was also an option, and another patient said to doubt between enucleation and PBT. Non-neutral framing typically occurred before the patients expressed a treatment preference (43/54, 80%). The final treatment decision corresponded to the preferred treatment expressed during the consultation for 49/54 (91%) patients. In 10 consultations in which patients did not express a preference, a treatment decision was made; for 9/10, this decision remained the final treatment choice. Ultimately, 73 (66%) patients received PBT, 34 (31%) underwent enucleation, and three (3%) received Ruthenium brachytherapy (Table 1).

### 3.4 Patient- and physician-perceived non-neutral framing

Ninety-two (84%) patients completed the iSHAREpatient, and the ocular oncologists completed 86 (78%) iSHAREphysician after seeing a participating patient. No significant correlations were observed between the presence of explicit non-neutral framing behaviour or the number of implicit non-neutral framing behaviours observed in the consultation, and the patient or ocular oncologist scores on how explicit the choice was presented, how well the options had been described, or whether patient preferences were taken into account from the participants’ perspectives (Table 2).

No statistically significant inter-oncologist variations were observed in the LMM sub-analysis regarding the correlation between the occurrence of explicit or implicit non-neutral framing behaviours, and how explicit the choice was presented, how well the options had been described, or whether patient preferences were taken into account in the decision-making from the participants’ perspectives (Supplemental Tables E.1 – E.3).

### 3.4 Patient satisfaction with decision-making

Eighty-one (74%) patients completed the SWD scale. The patients reported high satisfaction with their treatment decision (median = 30.0, range, 6 – 30), and more than half (48/81, 59%) scored the maximum score. No significant difference was found in patient satisfaction with decision-making if explicit non-neutral framing behaviour had occurred or not (*ρ* = -.114, 95% CI -0.33 – 0.11, p-value = 0.31). Also, the number of implicit non-neutral framing behaviours was not significantly related to patient satisfaction with decision-making (*ρ* = - .030,, 95% CI -0.25 – 0.19, p-value = 0.79).

## 4. Discussion

To the best of our knowledge, this study is the first to analyse neutrality of information provision about treatment options in ocular oncology, for consultations in which SDM is appropriate. Our study shows that in most consultations non-neutral framing behaviour was present. It occurred either explicitly, implicitly, or both, usually once, and it most often favoured eye-preserving therapy. Regardless of the occurrence of non-neutral framing behaviour, patients were satisfied with the decision-making process.

Explicit non-neutral framing occurred in about one-third of the consultations. It was noteworthy that in almost all consultations, the patient had not yet voiced or did not voice a treatment preference of what is important to them. Physicians should be mindful when stating treatment preferences for two reasons: it may falsely imply a medically superior option despite equipoise (as in this case regarding tumour control and metastasis risk), and it can unduly influence the patient’s own preference (24, 25). In hypothetical scenarios involving a clearly superior treatment option, patients have been found to prioritize the physician’s recommendation even when the recommended option does not maximize potential clinical outcomes (26). This suggests that a physician’s influence can even dominate objective treatment superiority. Patients may go along with the physician’s recommendation as it may feel counterintuitive from a patient’s perspective to oppose it (16, 27). However, a physician’s preference may not be useful for patients, as their priorities may differ (28). Ocular oncologists should therefore be careful to voice their own preference if they wish to involve patients meaningfully in treatment decision-making (19). Providing advice is appropriate on the condition that physicians have a good understanding of the individual patient’s preferences, incorporate what the patient prioritizes in their recommendation, and makes explicit how the recommendation fits patient’s preferences (12, 29). In that manner, the patient and ocular oncologist can evaluate if the ocular oncologist indeed considered what is most important to the patient in making the decision.

Three-quarters of the consultations contained implicit non-neutral framing. These results are comparable with previous findings suggesting that the occurrence of implicit non-neutral framing behaviour during consultations is common (14, 30-32). Implicit non-neutral framing by emphasizing treatments positively or negatively occurred often. This may stem from perceiving PBT as better given its potential to preserve the eye and thereby some vision, or the assumption that patients might regret not having attempted to preserve visual function. Enucleation was often framed as “serious”, probably because it entails certain loss of vision. Regardless of the reason for emphasizing a treatment, any assumption must be verified with the patient. In addition, ocular oncologists frequently shared extreme instead of nuanced patients’ experiences when describing treatments. Extreme examples are unlikely to reflect most patients, and fail to show how patients make the trade-offs between treatment options. Also, ocular oncologists often emphasized the benefits or harms of the treatment options. Within the same consultation, the benefits of one treatment option were often weighed against the disadvantages of the other, or only benefits or disadvantages were communicated. In either case, unbalanced presentation of advantages and disadvantages of possible treatments makes it more difficult for patients to weigh all relevant information. Lastly, we observed instances in which implicit non-neutral framing behaviour occurred toward both treatment options. This may neutralise the impact of wording on shaping a patient’s preference. Regardless, providing factual descriptions of what the treatment would mean for the patient allows them better to evaluate options in light of their personal circumstances and views.

Non-neutral framing behaviour occurred in most consultations, and was not associated with how the participants perceived the presentation of treatment choice, how equal the benefits and side-effects of the treatment options had been explained, or the extent to which patient’s preferences had been incorporated in the treatment decision. Possibly, some patients expect their ocular oncologist to explicitly recommend a treatment (33, 34). Patients may further not notice non-neutral framing because other aspects of the conversation take precedence, such as ocular oncologist’s empathy, approachability, and practical support. Furthermore, non-verbal communication was not accounted for in this study and non-verbal cues, such as the vocal immediacy or expressions of confirmation, may mask non-neutral framing behaviour. Indeed, in simulated consultations patients report higher satisfaction with physicians who smile more or make more eye contact (35). Moreover, patients may not have had sufficient time to reflect on the consultation when they filled in the iSHARE questionnaire, especially as they just had received their cancer diagnosis. The diagnosis likely caused patients to experience emotional stress, leaving little room for reflection on the decision-making process (36). The lack of association between non-neutral framing and how ocular oncologists perceived the decision-making process may be related to physicians’ limited ability to accurately assess their own communication skills (37). Moreover, how they frame information is often unintentional and routinized rather than deliberate (38). Additionally, the consultations under study were time-pressured. Evidence suggests that this may push physicians to frame information about treatment options favourably toward a specific option, to ensure clinical efficiency (39). Scheduling more time for these consultations could help reduce this tendency (40, 41). Of note, PBT and enucleation have comparable oncological outcomes, yet ocular oncologists may not find them equally medically fitting for a particular patient, for example due to the risk of (post-operative) complications given particular tumour dimensions. Explaining the clinical rationale for preferring one treatment is then appropriate and fosters SDM. When ocular oncologists justified their preference, mentioning their preference was not classified as explicit framing behaviour. Lastly, the LMM analysis showed no significant differences between the ocular oncologists’ perceptions of the decision-making process, and the use of explicit and implicit non-neutral framing behaviour. This points to a homogenous pattern in the study sample.

## 5. Strengths and limitations

A significant strength of this study is the relatively large cohort for a rare malignancy. The response rate for the questionnaires in this study was also substantial.

There were some limitations to the study. First, the audio recordings did not capture interactions between the ocular oncologist and the patient prior to the consultation, including intake and diagnostic testing. Ocular oncologists may have acquired personal or clinical information that was not repeated during the recorded consultation. This may have led to overestimating explicit non-neutral framing; possibly, ocular oncologists mentioned a preferred treatment that was in fact related to a preference of the patient’s individual characteristics, and 2) providing an evaluation of a treatment without providing a medically substantive clarification alongside. Yet, the recorded consultations were the formal encounters in which the final diagnosis was shared and treatment options were discussed, and therefore the first time at which patients could develop an informed treatment preference.

Second, selection bias may have occurred. Possibly, ocular oncologists may not have asked eligible patients for consent to audio record the consultation if they anticipated the consultation to be difficult or sensitive. Thereby, patients who may be more susceptible to the effects of non-neutral framing behaviours, or who might experience greater difficulty in determining their treatment preference could be underrepresented in the sample.

## 6. Conclusion

This study demonstrates that ocular oncologists frequently display one or more non-neutral framing behaviours in consultations, most often implicitly, and primarily favouring proton beam therapy. Patients and ocular oncologists did not seem to notice the framing behaviour, and non-neutral framing was not associated with patient satisfaction regarding decision-making. Neutral information provision is not fully feasible, but striving to minimize subjectivity when providing treatment information is desirable. Non-neutral framing can potentially influence patients’ preferences in ways that do not genuinely reflect their goals and priorities. More research is required to determine to what extent non-neutral framing impacts treatment decision-making.

## Data Availability

All data produced in the present work are contained in the manuscript.

## Acknowledgements

We would like to thank Joke Katerberg and Susanne Ravensbergen for helping with patient recruitment. We would also like to thank Jan-Willem Beenakker for his critical eye during our meetings about this study.

