## Supplemental Table for "Shared decision-making about uveal melanoma treatment in the Netherlands: non-neutral framing of medical information"

**Supplementary data**

**Supplemental Table A.** Definition of implicit non-neutral framing behaviors based on Engelhardt et al*.

| **Implicit non-neutral framing behavior** | **Definition** |
|---|---|
| From mild to more serious treatment - a gradual decision | Presenting the various treatment strategies in such a way that the proposed course of treatment seems to be the least aggressive or invasive, and consequently more appealing (or vice versa). |
| Deterring vs encouraging: *using others as an example* | Using other patients' frightening or hopeful stories as examples to convince patients to choose the course of treatment favoured by the physician. |
| Selectively presenting side-effects and/or benefits | Emphasizing either the benefits or harms of the treatment that was (un)favoured by the physician. |
| Emphasizing benefits or side-effects of treatment | Emphasizing the magnitude of the treatment effect and minimizes the side-effects or vice versa. |
| Presenting side-effects after final treatment decision had been made | Presenting the side-effects of a treatment after a decision on whether to start treatment or not was made. |
| Selectively presenting risks of side-effects | Focusing on only the most common and the least threatening side-effects. |
| Minimizing the treatment's impact | Downplaying the impact of the treatment. |
| Making assertions about the patient's personality or health status | Making assertions about what the patient could or could not handle, her ability to persevere and priorities in life, to steer her towards undergoing or foregoing treatment. |
| Presenting treatment as an authorized 'we' decision | Presenting the treatment as an authorized decision based on consensus amongst experts. |
| Presenting treatment as an authorized decision based on 'the guideline' | Presenting the treatment as an authorized decision based on guideline recommendation for patients with their personal (e.g. age) and/or disease characteristics (e.g. tumor size) |
| Travel distance** | Using travel distance in such a way that the course of treatment seems less or more appealing. |

***** *Engelhardt EG, Pieterse AH, van der Hout A, de Haes HJ, Kroep JR, Quarles van Ufford-Mannesse P, et al. Use of implicit persuasion in decision making about adjuvant cancer treatment: A potential barrier to shared decision making. Eur J Cancer. 2016;66:55-66.*
Some implicit behaviours from the above study were excluded from this study.
** This behavior was not part of the original coding scheme of Engelhardt et al.

**Supplementary data**

**Supplemental Table B.** iSHAREpatient [a, b]

<table>
<tr><td rowspan="8">Medical information</td><td colspan="6">1. The doctor explained what the advantages of treatment options are</td></tr>
<tr><td>☐ Not at all</td><td>☐ Hardly</td><td>☐ A little</td><td>☐ For a large part</td><td>☐ Almost completely</td><td>☐ Completely</td></tr>
<tr><td colspan="6">2. The doctor explained what the disadvantages of the treatment options are</td></tr>
<tr><td colspan="6">3. The doctor explained the advantages and disadvantages of each treatment option equally well</td></tr>
<tr><td colspan="6">4. The doctor checked whether I understood the advantages of the treatment options</td></tr>
<tr><td colspan="6">5. The doctor checked whether I understood the disadvantages of the treatment options</td></tr>
<tr><td colspan="6">6. The doctor told me how the treatment options differ from each other</td></tr>
<tr><td colspan="6">7. I asked questions about the treatment options</td></tr>
<tr><td rowspan="2">Choice awareness</td><td colspan="6">8. At the beginning of the conversation, the doctor said that there was a choice with regard to my treatment</td></tr>
<tr><td colspan="6">9. The doctor said that it matters what I think is important</td></tr>
<tr><td rowspan="2">Decision</td><td colspan="6">15. The decision takes into account what I consider to be important</td></tr>
<tr><td colspan="6">16. The doctor has discussed with me what I need in order to weigh up the advantages and disadvantages of the treatment options</td></tr>
</table>

[a] Bomhof-Roordink H, Gartner FR, van Duijn-Bakker N, van der Weijden T, Stiggelbout AM, Pieterse AH. Measuring shared decision making in oncology: Development and first testing of the iSHAREpatient and iSHAREphysician questionnaires. Health Expect. 2020;23(2):496-508.

[b] Following this study, the order of the items for both iSHARE questionnaires have been adjusted: items 8 and 9 are now items 1 and 2; items 10 and 13 have been reversed.

**Supplementary data**

**Supplemental Table C.** iSHAREphysician [a, b]

<table>
<tr><td rowspan="8">Medical information</td><td colspan="6">1. I explained what the advantages of treatment options are</td></tr>
<tr><td>☐ Not at all</td><td>☐ Hardly</td><td>☐ A little</td><td>☐ For a large part</td><td>☐ Almost completely</td><td>☐ Completely</td></tr>
<tr><td colspan="6">2. I explained what the disadvantages of the treatment options are</td></tr>
<tr><td colspan="6">3. I explained the advantages and disadvantages of each treatment option equally well</td></tr>
<tr><td colspan="6">4. I checked whether I understood the advantages of the treatment options</td></tr>
<tr><td colspan="6">5. I checked whether I understood the disadvantages of the treatment options</td></tr>
<tr><td colspan="6">6. I told me how the treatment options differ from each other</td></tr>
<tr><td colspan="6">7. The patient asked questions about the treatment options</td></tr>
<tr><td rowspan="2">Choice awareness</td><td colspan="6">8. At the beginning of the conversation, I said that there was a choice with regard to my treatment</td></tr>
<tr><td colspan="6">9. I said that it matters what I think is important</td></tr>
<tr><td rowspan="2">Decision</td><td colspan="6">10. The decision takes into account what the patient considers to be important</td></tr>
<tr><td colspan="6">11. I discussed with me what I need in order to weigh up the advantages and disadvantages of the treatment options</td></tr>
</table>

[a] Bomhof-Roordink H, Gartner FR, van Duijn-Bakker N, van der Weijden T, Stiggelbout AM, Pieterse AH. Measuring shared decision making in oncology: Development and first testing of the iSHAREpatient and iSHAREphysician questionnaires. Health Expect. 2020;23(2):496-508.

[b] Following this study, the order of the items for both iSHARE questionnaires have been adjusted: items 8 and 9 are now items 1 and 2; items 10 and 13 have been reversed.

**Supplementary data**

**Supplemental Table D.** Examples of quotes of implicit non-neutral framing behaviour

| | Example quote | In favor of treatment |
|---|---|---|
| From mild to more serious treatment - a gradual decision | We have listed them in order of attractiveness. | (the option that was mentioned first varied) |
| Deterring vs encouraging: using others as an example | And another patient says: a tumour in my body, it has to be removed today. I don't want anything to do with it anymore. It has to go, gone. I want that thing gone, gone, gone. | Enucleation |
| Selectively presenting risks of side-effects | The most important benefit for you is that you can keep your eye cosmetically, but with a whole lot of hassle involved. | Proton beam therapy |
| Emphasizing benefits or side-effects of treatment | In terms of vision, your eyesight will not improve from this. In fact, it could even get worse. […] As I mentioned before, when it comes to vision, you should know that this will not make it any better. | Enucleation |
| Presenting side-effects after final treatment decision had been made | **Patient:** Well, I would suggest that I still have some eyesight.<br>**Physician:** Yes, exactly. So then we'll use proton radiation.<br>**Patient:** Yes, yes.<br>**Physician:** Exactly. It does mean that you'll have to come here a bit more often.<br>**Patient:** Yes.<br>**Physician:** So you will have to travel back and forth. | Enucleation |
| Minimizing the treatment's impact | [*gives information about enucleation*].. An external prosthesis that is held in place under the eyelids, so all that's not complicated. | Enucleation |
| Making assertions about the patient's personality or health status | And usually, I think that you, for the proton treatment, you have to do quite a bit. MRI scans have to be done, and quite a lot of things need to happen. But looking at you now, I feel you can handle it. | Proton beam therapy |
| Presenting treatment as an authorized 'we' decision | But if you compare it to each other, it is quite intensive, and we really only recommend it for people who are very motivated and therefore willing to travel back and forth all the time. | Proton beam therapy |
| Presenting treatment as an authorized decision based on 'the guideline' | There is also another, something else we could do, and we only do that in patients if the tumour is very large and if it is the patient's explicit wish — which means removing the entire eye. | Enucleation |
| Travel distance | Proton treatment is a bit more complicated, because we need to do an MRI before the surgery and after the surgery, you'll have regular check-ups for which you'll need to come here. Fortunately, you don't live too far from here. | Proton beam therapy |

**Supplementary data**

**Supplemental Table E.1.** Correlations between Dimension 1 of the iSHARE questionnaires and manifestations of non-neutral framing, by ocular oncologist (N=4)

| iSHAREphysician | | | | | |
|---|---|---|---|---|---|
| | | A | B | C | D |
| | Dimension mean score | 4.4 | 3.5 | 3.3 | 4.0 |
| Explicit non-neutral framing behaviour (yes/no) | Estimate | 0.360 | 1.071 | 1.687 | 0.447 |
| | SE | 0.097 | 0.426 | 0.616 | 0.158 |
| | *P*-value | 0.986 | | | |
| Total number of implicit non-neutral framing behaviours | Dimension score | 4.4 | 3.5 | 3.3 | 4.0 |
| | Estimate coefficient | 0.337 | 1.051 | 1.820 | 0.429 |
| | SE | 0.087 | 0.416 | 0.670 | 0.149 |
| | *P*-value | 0.151 | | | |
| iSHAREpatient | | | | | |
| | Dimension mean score | 4.6 | 4.7 | 5.0 | 4.9 |
| Explicit non-neutral framing behaviour (yes/no) | Estimate | 0.790 | 0.288 | 0.014 | 0.206 |
| | SE | 0.187 | 0.098 | 0.006 | 0.066 |
| | *P*-value | 0.264 | | | |
| Total number implicit non-neutral framing behaviour | Dimension mean score | 4.6 | 4.7 | 5.0 | 4.9 |
| | Estimate coefficient | 0.791 | 0.263 | 0.017 | 0.188 |
| | SE | 0.187 | 0.089 | 0.006 | 0.060 |
| | *P*-value | 0.133 | | | |

**Supplementary data**

**Supplemental Table E.2.** Correlations between Dimension 2 of the iSHARE questionnaires and manifestations of non-neutral framing, by ocular oncologist (N=4)

| | | | | | |
|---|---|---|---|---|---|
| iSHAREphysician | | | | | |
| | | Physician A | Physician B | Physician C | Physician D |
| Explicit non-neutral framing behaviour (yes/no) | Dimension mean score | 3.929 | 3.020 | 2.929 | 3.330 |
| | Estimate | 1.006 | 0.218 | 0.649 | 0.280 |
| | SE | 0.287 | 0.088 | 0.244 | 0.101 |
| | *P*-value | 0.594 | | | |
| Total number implicit non-neutral framing behaviour | Dimension mean score | 3.9 | 3.0 | 2.7 | 2.9 |
| | Estimate coefficient | 0.932 | 0.237 | 0.703 | 0.285 |
| | SE | 0.286 | 0.095 | 0.282 | 0.104 |
| | *P*-value | 0.508 | | | |
| iSHAREpatient | | | | | |
| Explicit non-neutral framing behaviour (yes/no) | Dimension mean score | 4.2 | 4.3 | 4.6 | 4.7 |
| | Estimate | 1.775 | 0.591 | 0.717 | 0.482 |
| | SE | 0.417 | 0.201 | 0.250 | 0.162 |
| | *P*-value | 0.605 | | | |
| Total number implicit non-neutral framing behaviour | Dimension mean score | 4.2 | 4.3 | 4.6 | 4.4 |
| | Estimate coefficient | 1.783 | 0.571 | 0.728 | 0.481 |
| | SE | 0.420 | 0.196 | 0.251 | 0.163 |
| | *P*-value | 0.475 | | | |

**Supplementary data**

**Supplemental Table E.3.** Correlations between Dimension 6 of the iSHARE questionnaires and manifestations of non-neutral framing, by ocular oncologist (N=4)

| iSHAREphysician | | | | | |
|---|---|---|---|---|---|
| | | Physician A | Physician B | Physician C | Physician D |
| Explicit non-neutral framing behaviour (yes/no) | Dimension mean score | 4.1 | 3.3 | 2.9 | 3.2 |
| | Estimate | 0.929 | 0.513 | 0.973 | 0.633 |
| | SE | 0.283 | 0.193 | 0.380 | 0.232 |
| | *P*-value | 0.128 | | | |
| Total number implicit non-neutral framing behaviour | Dimension mean score | 4.1 | 3.3 | 2.9 | 3.2 |
| | Estimate coefficient | 0.787 | 0.589 | 1.133 | 0.654 |
| | SE | 0.251 | 0.233 | 0.469 | 0.232 |
| | *P*-value | 0.073 | | | |
| iSHAREpatient | | | | | |
| Explicit non-neutral framing behaviour (yes/no) | Dimension mean score | 4.7 | 4.7 | 4.6 | 4.6 |
| | Estimate | 0.662 | 0.652 | 1.397 | 0.366 |
| | SE | 0.160 | 0.220 | 0.482 | 0.118 |
| | *P*-value | 0.425 | | | |
| Total number implicit non-neutral framing behaviour | Dimension mean score | 4.8 | 4.7 | 4.6 | 4.6 |
| | Estimate coefficient | 0.649 | 0.684 | 1.439 | 0.364 |
| | SE | 0.160 | 0.230 | 0.494 | 0.119 |
| | *P*-value | 0.833 | | | |
